# Disrupted cerebellar structural connectome in spinocerebellar ataxia type 3 and its association with transcriptional profiles

**DOI:** 10.1101/2023.09.13.23295487

**Authors:** Xinyi Dong, Bing Liu, Weijie Huang, Haojie Chen, Yunhao Zhang, Amir Shmuel, Zeshan Yao, Guolin Ma, Ni Shu

**Affiliations:** State Key Laboratory of Cognitive Neuroscience and Learning and IDG/McGovern Institute for Brain Research, Beijing Normal University, Beijing 100875, China; BABRI Centre, Beijing Normal University, Beijing 100875, China; Beijing Key Laboratory of Brain Imaging and Connectomics, Beijing Normal University, Beijing 100875, China; Department of Radiology, Shandong Provincial Hospital Affiliated to Shandong First Medical University, Jinan, Shandong Province, 250021, China; Department of Radiology, China-Japan Friendship Hospital, Beijing 100029, China; Department of Systems Science, Beijing Normal University, Beijing 100875, China; State Key Laboratory of Multimodal Artificial Intelligence Systems, Institute of Automation, CAS, Beijing 100190, China; McConnell Brain Imaging Centre, Montreal Neurological Institute, McGill University, Montreal, QC, Canada; Departments of Neurology and Neurosurgery, Physiology, and Biomedical Engineering, McGill University, Montreal, QC, Canada; Institute of Biomedical Engineering, Jingjinji National Center of Technology Innovation, Beijing 100094, China

**Author notes:** Correspondence to: Ni Shu State Key Laboratory of Cognitive Neuroscience and Learning and IDG/McGovern Institute for Brain Research, Beijing Normal University Beijing 100875, China Correspondence may also be addressed to: Guolin Ma Department of Radiology, China-Japan Friendship Hospital Beijing 100029, China. ^†^These authors contributed equally to this work.

**Keywords:** spinocerebellar ataxia type 3, cerebellar structural connectome, white matter, gene expression, topology

## Abstract

Spinocerebellar ataxia type 3 (SCA3) is primarily characterized by progressive cerebellar degeneration, including gray matter atrophy and disrupted anatomical and functional connectivity of the cerebellum. The alterations of topological organization of cerebellar white matter structural network in SCA3 and the underlying neurobiological mechanism remain unknown. Using a cohort of 20 patients with SCA3 and 20 healthy controls, we constructed cerebellar structural networks from diffusion magnetic resonance imaging and investigated alterations of topological organization. Then we mapped the alterations with transcriptome data from the Allen Human Brain Atlas to identify possible biological mechanisms for regional selective vulnerability to white matter damage. Compared with healthy controls, decreased global and nodal efficiency, and widely distributed decreased edge strength were observed in SCA3 patients. The regions with decreased nodal global efficiency were mainly located in cerebellar anterior lobe, and the genes express higher in these regions were significantly enriched in synapse-related biological processes. The regions with decreased nodal local efficiency were mainly located in cerebellar posterior lobe, and the higher gene expression in these regions were significantly enriched in metabolic biological processes. Similar hub distributions were found in two groups of subjects, whereas the strengths of rich-club and feeder connections were lower in SCA3 patients. Moreover, strength of the inter-module connections was lower in SCA3 group and negatively correlated with SARA score, ICARS score, and CAG repeat number. These findings suggest a mechanism of white matter vulnerability and a potential image biomarker for the disease severity, providing insights into neurodegeneration and pathogenesis in this disease.

## Introduction

Spinocerebellar ataxia type 3 (SCA3), also known as Machado-Joseph disease (MJD), is an inherited neurodegenerative disease caused by a CAG repeat expansion in the coding region of the ATXN3 gene for a polyglutamine (polyQ) stretch^1^ and is the most common subtype of hereditary ataxia^2^. The primary feature of SCA3 is progressive cerebellar degeneration, leading to symptoms including gait abnormalities, dysarthria, and abnormal eye movements^3^. In postmortem cerebellar tissue from SCA3 patients, reduced myelin staining and myelin basic protein has been discovered in cerebellar lobules^4,5^. Recent neuroimaging studies reported that SCA3 is accompanied by gray matter atrophy^6–9^, white matter abnormalities^10–12^ and functional disruption^6,13^. However, whether SCA3 patients exhibit alterations of brain connectivity and what is the neurobiological mechanism underlying the abnormalities remain to be elucidated.

Diffusion MRI (dMRI) has been extensively used to visualize white matter tracts non-invasively in vivo. Some dMRI studies have reported white mater microstructural abnormalities such as decreased fractional anisotropy (FA) and increased mean diffusivity (MD) in regions including the cerebellar hemispheres, cerebellar vermis, pons, dentate nucleus, basal ganglia, and thalamus of SCA3 patients^10–12,14,15^. Recent neuroimaging studies model the human brain as a network, referred to as a connectome^16^, to quantify the communication between regions of interest. The human brain connectome exhibits several remarkable topological attributes, such as small-worldness, rich-club organization, and modular structure in healthy population^17,18^. A recent study investigated alterations in the white matter (WM) structural motor network, and found global topological measures of the WM motor network to be a promising image biomarker for SCA3 disease severity^19^. However, the topological organization of cerebellar WM network hasn’t been studied systematically and how the cerebellar network alters in SCA3 patients is still unclear.

SCA3 is one of the most frequent polyQ diseases characterized by the formation of protein aggregates in neurons^20^. Pathological aggregates can affect other proteins and cellular components, progressively leading to the dysregulation and impairment of several cellular mechanisms^21,22^. Despite the identification of the causative mutation, the biological processes that lead to neurodegeneration observed in the disease are not yet fully understood^21^. The emergence of transcriptome-neuroimaging association study has made it possible to bridge the gap between the large-scale transcriptional profile and connectome topology^23,24^. Therefore, if SCA3 patients show abnormalities in WM structural network, the transcriptome-neuroimaging association study can identify related gene expression profiles so as to understand why some regions exhibit selective vulnerability^25^ to white matter damage. However, such study hasn’t been performed so far to explore the molecular genetic underpinnings of SCA3 disease.

To address these gaps, we used dMRI data and graph theory approach to investigate the topological alterations in the structural connectome in patients with SCA3 and established the association with the transcriptome profile to identify possible mechanisms for regional selective vulnerability to white matter damage in SCA3 disease. We hypothesized that: first, the topological attributes of cerebellar structural network are disrupted in patients with SCA3; second, regional gene expression profiles associated with regional SCA3-related alterations are involved in specific biological processes such as synapse-related functions.

## Materials and methods

### Participants

This study was approved by the Ethics Committee of China-Japan Friendship Hospital. Written informed consent were obtained from all subjects.

Twenty SCA3 patients (16 males and 4 females; mean age, 46.0±13.2 years; age range, 25-67 years) and twenty age, sex and education-matched healthy controls (HCs) (15 males and 5 females; mean age, 46.5±15.6 years; age range, 24-72 years) were enrolled. All subjects were right-handed. The inclusion criteria for SCA3 patients were: (1) diagnosed as SCA3 by expert neurologists according to the family history, neurological manifestations, genetic and molecular tests, and routine brain MRI findings; (2) age≥18 years; (3) right-handed. Patients who had contraindications to MRI examination or suffered from other neurological diseases were excluded from this study. The HCs were recruited from the local communities with age ≥18 years, right-handed, and had no history of any neuropsychiatric disorders. Demographics collection were performed before MR data acquisition on the scanning day. Meanwhile, the Scale for the Assessment and Rating of Ataxia (SARA)^26^ and the International Cooperative Ataxia Rating Scale (ICARS)^27^ scores were examined and recorded by trained neurologists to measure the disease severity of SCA3 patients.

### Image acquisition

MRI studies of all subjects were carried out on a 3.0 Tesla MR scanner (General Electric, Discovery MR750, Milwaukee, WI, United States) with an 8-channel head coil. Earplugs and paddings were provided to all subjects for noise reduction, and foam paddings were given to restrict head motion. During the MR studies, all subjects were asked to keep their eyes closed without falling asleep, relax and think nothing special.

The structural T1 weighted volumes acquisition and the diffusion weighted acquisition were included in the MRI sequences. Morphological T1 weighted volumetric sequence were based on the three-dimensional fast spoiled gradient-echo sequences (3D FSPGR) with the following parameters: repetition time (TR) = 6.7 ms, echo time (TE) = 2.9 ms, flip angle = 12°, slice thickness = 1 mm, field of view (FOV) = 256×256 mm^2^, matrix = 256×256, voxel size = 1×1×1 mm^3^. A single-shell diffusion weighted spin-echo echo-planar sequence (EPI) was used for dMRI data with 64 encoding directions (b = 1,000 s/mm^2^) and 8 b0 reference images (b = 0 s/mm^2^), TR = 8,028 ms, TE = 81.8 ms, flip angle = 90°, slice thickness = 2 mm, FOV = 240 × 240 mm^2^,matrix = 120 ×120, voxel size = 2×2×2 mm^3^.

### Image processing

Preprocessing of T1w and dMRI data was performed in QSIPrep^28^ (v.0.16.0RC3), which integrates the other existing packages such as FSL^29^, DSI Studio^30^, DIPY^31^, ANTs^32^ and MRtrix^33^.

T1w images were corrected for intensity non-uniformity using *N4BiasFieldCorrection* function and were skull-stripped to generate brain mask using *antsBrainExtraction.sh* in ANTs. Spatial normalization from T1w to MNI space was performed through nonlinear registration with *antsRegistration* function in ANTs. Brain tissue segmentation of cerebrospinal fluid (CSF), white-matter (WM) and gray-matter (GM) was performed on the brain-extracted T1w using *FAST* in FSL. For dMRI images, MP-PCA denoising with five-voxel window was implemented in MRtrix3’s *dwidenoise*. Then, B1 filed inhomogeneity was corrected with *dwibiascorrect* in MRtrix3. And the intensity of b=0 images is harmonized across scans. FSL’s *eddy* was used for head motion correction and eddy current correction. Finally, the b=0 template image was registered to the skull-stripped T1w image using ANTs.

### Cerebellar network construction

In order to construct cerebellar network, the preprocessing outputs were used to reconstruct white matter fiber tracts using a preconfigured reconstruction workflow provided by QSIprep (mrtrix_singleshell_ss3t). The workflow uses a constrained spherical deconvolution (CSD) algorithm named *ss3t_csd_beta1*^34^ to estimate Fibre orientations densities (FODs) for white matter using single shell (DTI) acquisitions. The white matter FODs are used for tractography using T1w segmentation by FSL’s *FAST* for anatomical constraints. The tractography is performed using MRtrix3’s *tckgen*, which uses the iFOD2 probabilistic tracking method to generate 10 million streamlines. Then, the weights of each streamline are calculated using spherical deconvolution informed filtering of tractograms 2 (SIFT2)^35^ method. To construct structural networks, the SUIT^36^ atlas was used to parcel the cerebellum into 34 regions defined as network nodes (Table 1), and Brainnetome^37^ atlas was used to parcel the cerebrum into 246 regions. The weight of streamlines connecting a pair of regions was defined as weight of network edge. Therefore, a weighted 34 × 34 cerebellar network and a weighted 246 × 246 cerebral network were constructed for each participant. The matrices of these networks remained unthresholded to stay consistent with recent studies^38,39^.

**Table 1.** Cerebellar regions of interest defined in the study.

| Region | Left/right/vermis | Label ID |
| --- | --- | --- |
| **Cerebellar cortex** |  |  |
| I_IV | Left, right | 1, 2 |
| V | Left, right | 3, 4 |
| VI | Left, vermis, right | 5, 6, 7 |
| CrusI | Left, vermis, right | 8, 9, 10 |
| CrusII | Left, vermis, right | 11, 12, 13 |
| VIIb | Left, vermis, right | 14, 15, 16 |
| VIIIa | Left, vermis, right | 17, 18, 19 |
| VIIIb | Left, vermis, right | 20, 21, 22 |
| IX | Left, vermis, right | 23, 24, 25 |
| X | Left, vermis, right | 26, 27, 28 |
| **Cerebellar nuclei** |  |  |
| Dentate | Left, right | 29, 30 |
| Interposed | Left, right | 31, 32 |
| Fastigial | Left, right | 33, 34 |

### Network analysis

To explore the topological organization of WM structural connectome, the graph metrics from the angles of network efficiency, hub distribution and modular parcellation were assessed.

### Network efficiency

To describe the topologic organization of the WM structural networks, various topological properties of a network from both global and nodal characteristics were evaluated. Global characteristics included global efficiency (𝐸_𝑔𝑙𝑜𝑏_), local efficiency (𝐸_𝑙𝑜𝑐_), clustering coefficient, shortest path length, and small-world parameters (γ, λ, σ). Regional characteristics included nodal global efficiency (𝑁𝐸_𝑔𝑙𝑜𝑏_), nodal local efficiency (𝑁𝐸_𝑙𝑜𝑐_), and degree centrality. Notably, 𝐸_𝑔𝑙𝑜𝑏_and 𝐸_𝑙𝑜𝑐_quantify integration and segregation of the brain network, respectively. The definitions of network efficiency^40^ are as follows.

The global efficiency of G measures the global efficiency of the parallel information transfer in the network, which is defined as:

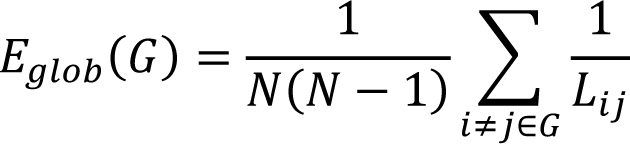

where 𝐿_𝑖𝑗_ is the shortest weighted path length between node 𝑖 and 𝑗 in 𝐺.

The local efficiency of 𝐺 shows how efficient the communication is among the first neighbors of the node 𝑖 when it is removed, is defined as:

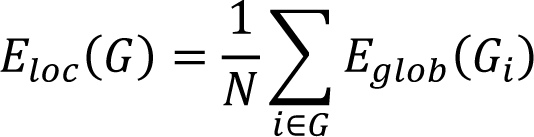

where 𝐺_𝑖_ is the subgraph composed of the nearest neighbors of node 𝑖.

The nodal global efficiency of one node measures the average shortest path length between a given node 𝑖 and all of the other nodes in the network, which is defined as:

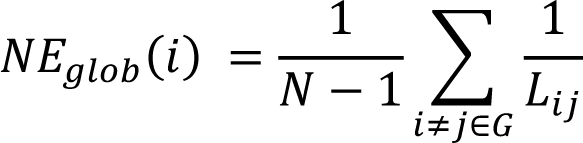

where 𝐿_𝑖𝑗_ is the shortest weighted path length between node 𝑖 and 𝑗 in 𝐺.

The nodal local efficiency is defined as:

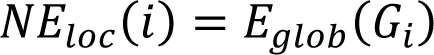

where 𝐺_𝑖_ is the subgraph composed of the nearest neighbors of node 𝑖.

### Hub distribution

The hub regions of WM structural network in each group were identified based on group-averaged backbone network. The backbone network was calculated by performing one-tailed sign test (P < 0.05, Bonferroni corrected) to save connections that were most consistent across subjects. Based on the backbone network, hub regions were defined as the nodes with degree centrality at least one standard deviation above the mean degree centrality across all regions. The regions were identified as hubs and non-hubs, and edges of the network were classified into rich-club connections, linking hubs; feeder connections, linking hubs and non-hubs, and local connections linking non-hubs. Finally, the sum strengths of rich-club, feeder, and local connections were separately calculated for each participant.

### Modular parcellation

To further explore the topological structure of the network, the brain network was parcelled into modules. The modularity Q(p) for a given partition p of the brain network quantifies the difference between the number of intra-module connections of actual network and that of random network^41^:

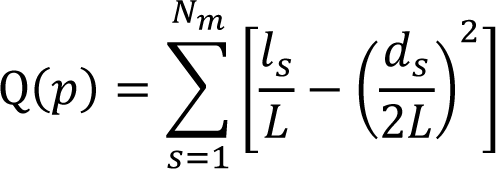

where 𝑁_𝑚_is the number of modules, 𝐿 is the number of connections in the network, 𝑙_𝑠_ is the number of connections between nodes in module 𝑠, and 𝑑_𝑠_is the sum of the degrees of the nodes in module s. Based on the backbone network of each group, the modular parcellation was performed to find a specific partition (p) which achieves the largest network modularity Q(p). Then, the sum strengths of intra-module and inter-module connections were calculated for each participant based on the partition within the group.

The calculation of global and local network properties and modular analysis were performed using GRETNA toolbox^42^ (http://www.nitrc.org/projects/gretna/). The results were visualized using BrainNet Viewer toolbox^43^ (http://www.nitrc.org/projects/bnv/).

### Statistical analysis

#### Group differences

Differences of demographics including age and education between SCA3 and HC group were examined with two-sample *t* tests. Difference of gender distribution was examined with 𝜒^2^ test. To examine between-group difference of global and nodal network properties, a general linear model (GLM) was performed with group as the main effect and gender, age, education as covariates. For nodal properties, false discovery rate (FDR) was used for multiple correction. For the network metrics with significant group differences, we performed a receiver operating characteristic curve analysis to evaluate the power of the metrics to discriminate SCA3 patients from healthy controls.

#### NBS analysis

To identify network edges that differ between two groups, network-based statistic (NBS) approach^44^ was implemented in GRETNA toolbox^42^. Firstly, a backbone which was intersection of SCA3 and HC backbone matrices was generated and masked to all subjects’ network matrices. Then, the same GLM analysis with gender, age and education as covariates was performed on all edges. A threshold of P < 0.000089 (0.05/34/33×2, Bonferroni correction) was used to select supra-threshold edges. Finally, among the set of supra-threshold edges, connected components and its significance were identified using a nonparametric permutation approach (10000 permutations). A threshold of P < 0.05 was used to select supra-threshold edges forming the component for each permutation.

#### Clinical correlation

To explore the power of network properties explaining clinical performance for SCA3 patients, we performed partial correlation analyses between network metrics and clinical scores within the SCA3 group controlling the effects of gender, age and education.

### Transcriptome-connectome association analysis

#### Estimation of regional gene expression

Regional gene expression profiles were obtained from the Allen Human Brain Atlas (AHBA)^24^ provided by the Allen Institute for Brain Science (AIBS) (http://human.brain-map.org, RRID: SCR_007416). The human brain tissue samples from this atlas were collected from the post-mortem brains of six adult donors (mean age: 42.5 years, range: 24–57 years, 1 female), including two complete brains and four left hemispheres. Preprocessing of the gene expression data was implemented in the *abagen* toolbox^45^. Tissue samples were assigned to their closest regions in SUIT atlas and samples with distance greater than 2 mm to any of the 34 regions were excluded. The procedure resulted in samples covering 28 regions. Expression values within each brain across regions were normalized with a scaled robust sigmoid function^46^. Finally, regional gene expression values were averaged across donors forming a 28 × 15633 regional transcription matrix X (Fig. 4A).

#### Partial least squares regression

Partial least square (PLS) regression was used to explore the association between the transcriptional profiles and alterations of regional network properties in SCA3. PLS regression is an unsupervised multivariate analysis producing components from X that have maximum covariance with Y (Fig. 4A). Y represents nodal global and local efficiency alterations of 28 regions, separately (28 × 1 vector). The analysis was performed using the *PLSRegression* function in Python package scikit-learn (https://scikit-learn.org/stable/index.html) with number of component set as 1.

The weights of all genes were obtained by PLS regression. To obtain reliable estimation of the weights, PLS regression was performed with bootstrap resampling resulting in distribution of each weight. The final weights were calculated with true weights divided by standard errors of the distributions.

#### Gene set enrichment analysis

We used a list of prioritized genes as input for gene set enrichment analysis implemented with GENE2FUNC function in FUMA^47^ online platform (https://fuma.ctglab.nl/). The genes were tested against Gene Ontology (GO) gene sets obtained from MsigDB^48^ on the basis of hypergeometric test. The set of background genes is protein-coding genes. Benjamini-Hochberg (FDR) multiple testing correction was performed per data source of tested gene sets. Gene sets with adjusted P-value less than 0.05 and the number of overlapping genes with gene-sets over 2 were reported. And the significant GO terms were visualized with REViGO^49^ online tool (http://revigo.irb.hr) to summarize them by removing redundant terms. Then, genes were tested against differentially expressed gene sets (DEG; genes significantly more or less expressing in a given tissue compared to others) across 54 tissue types obtained from GTEx v8^50^ to evaluate if the prioritized genes are overrepresented in specific tissue types.

## Results

### Disrupted network efficiency in SCA3

There were no significant differences in gender, age, and years of education between SCA3 and HC groups (Table 2). Both groups showed no small-world organization (σ < 1) of the cerebellar WM networks. In the cerebellar network, SCA3 have significantly less global efficiency (t = -3.16, P = 0.003), less local efficiency (t = -3.50, P = 0.001), less clustering coefficient (t = -2.68, P = 0.011) and more shortest path length (t = 3.06, P = 0.004) (Fig. 1A) as compared to HC groups. As for cerebral network, all global properties show no difference between two groups (all p > 0.05).

**Table 2.** Participant demographics and clinical characteristics.

| Measure | Healthy controls (n=20) | SCA3 patients (n=20) | Statistical value | P value |
| --- | --- | --- | --- | --- |
| Gender (M/F) | 15/5 | 16/4 | 0.37 | 0.714 |
| Age, years | 46.5 ± 15.6 | 46.0 ± 13.2 | -0.11 | 0.913 |
| Education, years | 12.6 ± 3.9 | 12.5 ± 4.2 | -0.12 | 0.908 |
| Disease duration, years |  | 9.3 ± 9.1 |  |  |
| SARA |  | 17.0 ± 10.8 |  |  |
| ICARS |  | 44.9 ± 24.8 |  |  |

Average values are reported as mean ± SD.
SCA3 = Spinocerebellar ataxias type 3; M = male; F = female; SARA = Scale for the Assessment and Rating of Ataxia; ICARS = International Cooperative Ataxia Rating Scale.

**Figure 1.**
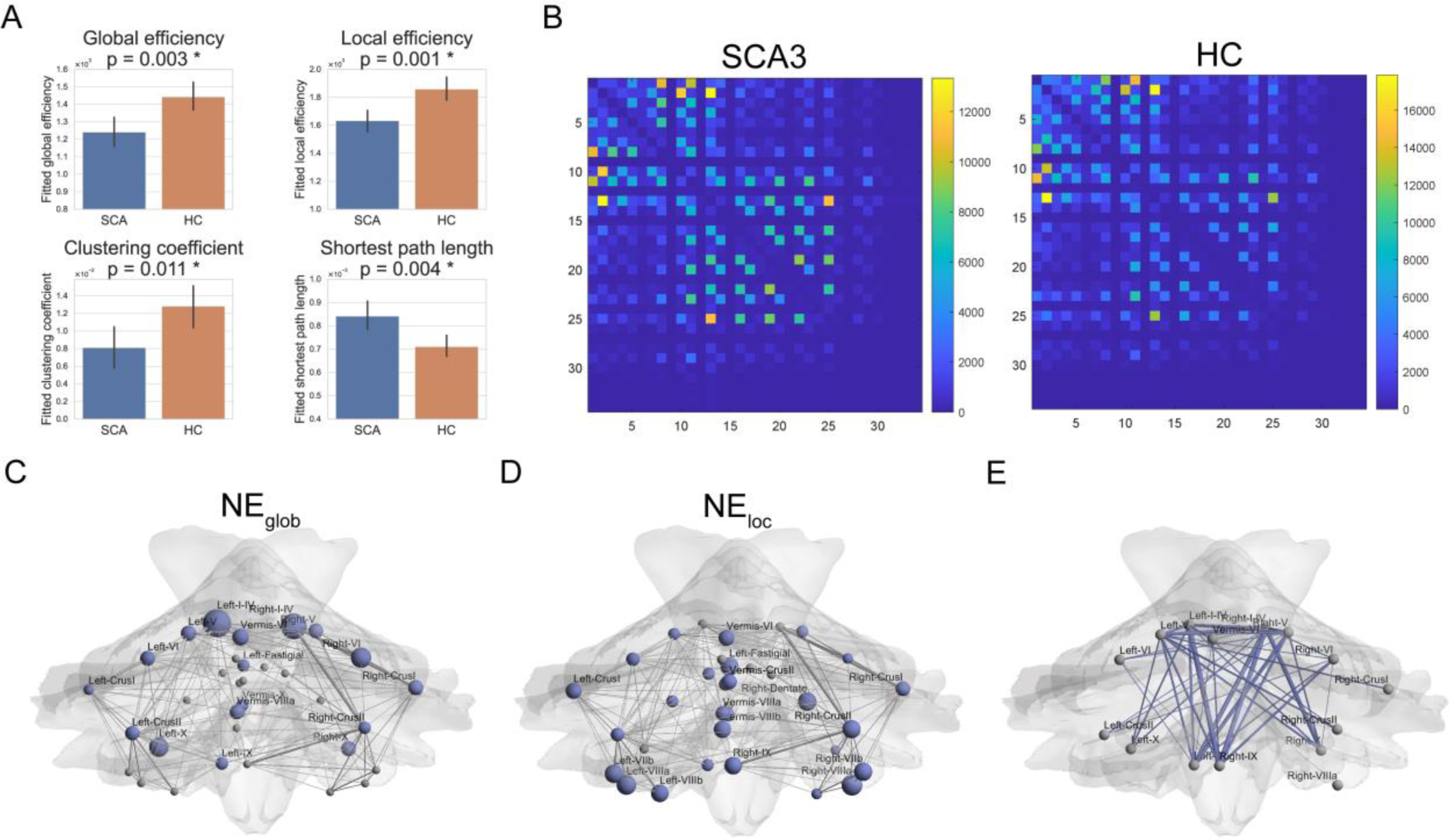
Topological differences between SCA3 patients and healthy controls on the global, nodal and edge level. (**A**) Group differences in the global properties of cerebellar WM structural networks. The fitted values indicate the residuals of the measures after removing the effects of gender, age and years of education. The bars and error bars represent the mean and standard deviations of fitted values, respectively. (**B**) The cerebellar backbone network of SCA3 participants (left) and healthy participants (right). (**C**) The distribution of the cerebellum regions with significantly lower nodal global efficiency in the SCA3 group. (**D**) The distribution of the cerebellum regions with significantly lower nodal local efficiency in the SCA3 group. The regions with significant group differences (P < 0.05, false discovery rate corrected) were colored in blue. The node size indicates the significance of the between-group differences. The edges were obtained by the backbone network of healthy participants with a sparsity of 20% for visualization. (**E**) A subnetwork with lower connection strength in the SCA3 group compared with the healthy controls revealed by network-based statistic. The edge size indicates the significance of the between-group differences.

We identified regions with less nodal global efficiency (Fig. 1C) and less nodal local efficiency (Fig. 1D) in the SCA3 group (P < 0.05, FDR corrected). The regions which have significant between-group difference of 𝑁𝐸_𝑔𝑙𝑜𝑏_mainly included bilateral I-IV, bilateral V, bilateral VI, bilateral CrusII, bilateral X, Vermis VI. Whereas the regions which have significant between-group difference of 𝑁𝐸_𝑙𝑜𝑐_mainly included bilateral VIIIa, right Dentate, bilateral VIIb, and right CrusII.

### Decreased connection strength in SCA3

The backbone network of SCA3 group and healthy control group is respectively shown in Fig. 1B. NBS analysis revealed a single connected subnetwork (component) with 15 nodes and 40 connections, which showed lower connection strength in the SCA3 group compared with the control participants (P < 0.05, FDR corrected; Fig. 1E). The disrupted structural connectivity was mainly distributed in the connections among regions like bilateral I-IV, bilateral V, bilateral X, and bilateral IX.

### Rich-club organization of WM Network in SCA3

Similar hub distribution was found between the SCA3 and HC groups, mainly located in the right-crusII, left-crusII, right-I-IV and left-crusI (Fig. 2A). However, the region left-I-IV was identified only in HC group, and the region right-IX was identified only in SCA3 group.

**Figure 2.**
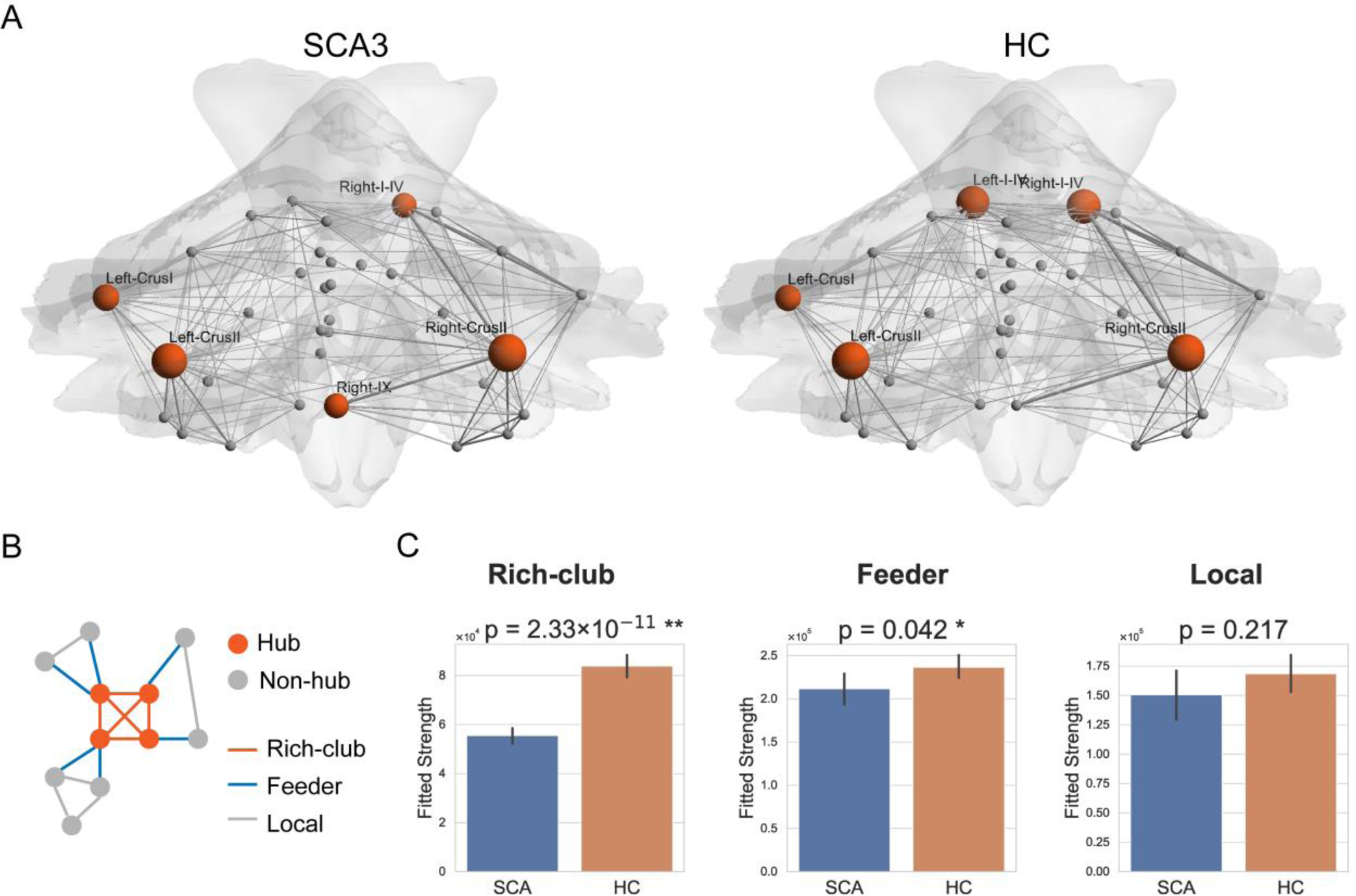
Rich-club organization of the cerebellar WM networks in SCA3 and healthy control groups. (**A**) The distributions of hub regions in two groups. The nodes with orange color represent the hub regions and the node size indicates the degree centrality of regions. The edges were obtained by the backbone network of each group with a sparsity of 20% for visualization. (**B**) An illustration of rich-club, feeder and local connections. (**C**) Group differences in the rich-club, feeder and local connection strengths. The fitted values indicate the residuals of the measures after removing the effects of gender, age and years of education. The bars and error bars represent the mean and standard deviations of fitted values, respectively.

As for the connections classified into rich-club, feeder, and local connections, the SCA3 group showed significantly lower strength of the rich-club connections (t = -9.62, P = 2.33×10^-11^), lower strength of the feeder connections (t = -2.11, P = 0.042), while no significant difference was found for the local connections (t = -1.26, P = 0.217) (Fig. 2C).

### Modular structure of WM Network in SCA3

We performed modular analysis based on the backbone network of SCA and HC group. The modularity value is 0.35 for SCA group and 0.29 for HC group. Two modules were identified in both groups, where module I mostly concluded regions in left hemisphere and module II mostly concluded regions in right hemisphere (Fig. 3A). Then, the edge strength of both intra-module and inter-module across two modules were calculated for each subject. SCA3 group showed significantly lower strength of the inter-module connections (t = -7.69, P = 5.06×10^-9^), lower strength of intra-module I connections (t = -2.14, P = 0.039), while no significant difference was found for strength of intra-module II connections (t = -1.91, P = 0.850) (Fig. 3C). The receiver operating characteristic curve analysis revealed that inter-module connection strength exhibited the highest area under the curve value of 0.96 for the discrimination between SCA3 and healthy control participants (Fig. 3B).

**Figure 3.**
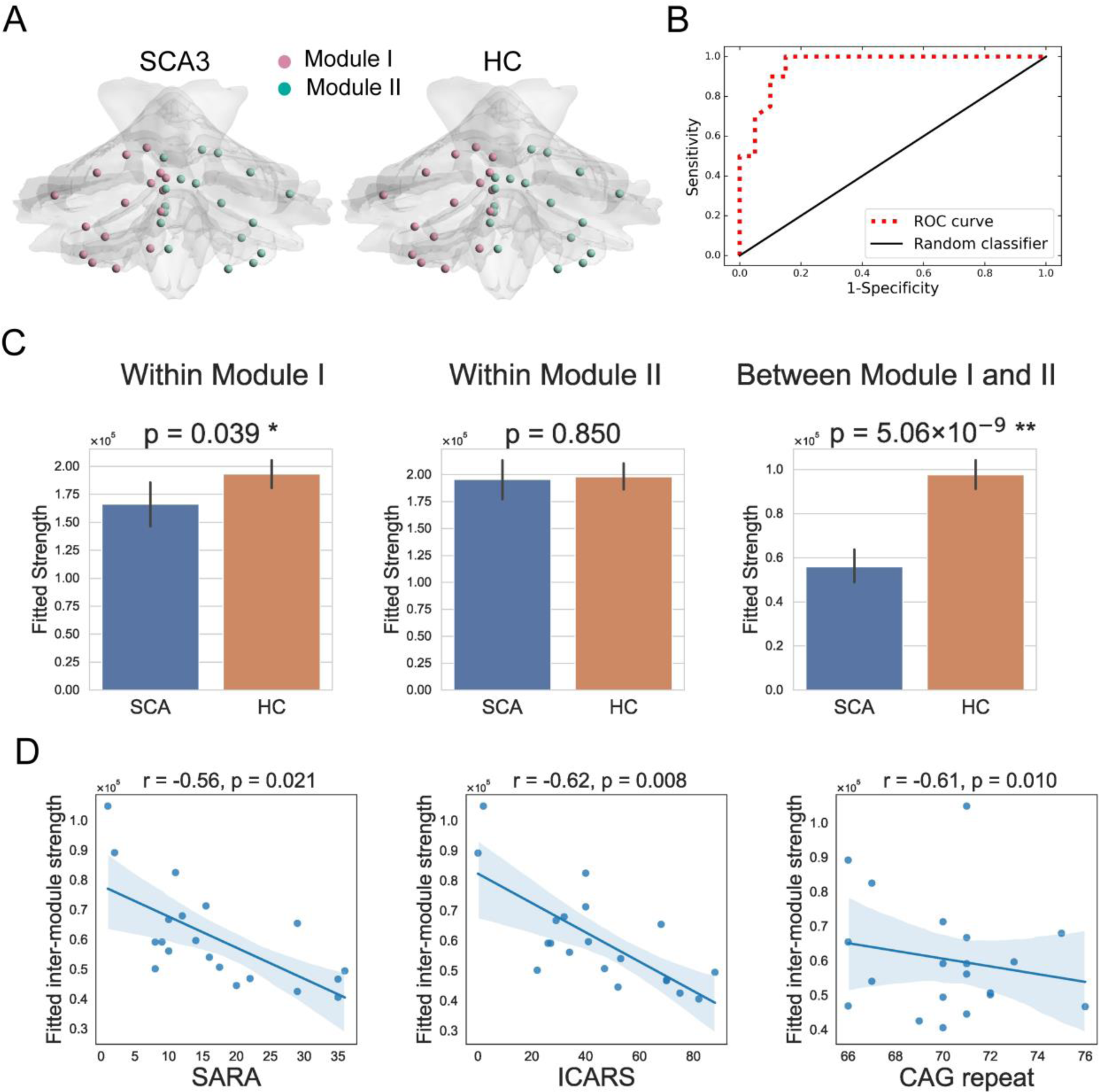
Modular structure of the cerebellar WM networks in SCA3 and healthy control groups. (**A**) Modular parcellation identifies two modules based on the backbone network of each group. (**B**) The receiver operating characteristic (ROC) curve of the inter-module connection strength exhibits good performance (area under the curve, 0.96) to differentiate SCA3 participants from healthy control participants. (**C**) Group differences in the inter-module and intra-module connection strengths. The bars and error bars represent the mean and standard deviations of fitted values, respectively. (**D**) Scatterplots show the significantly negative correlations between inter-module connection strength and clinical scores of SARA score, ICARS score and CAG repeat number in SCA3 patients. The fitted values indicate the residuals of the measures after removing the effects of gender, age and years of education.

### Relationships between network attributes and clinical scores

Within the SCA3 group, strength of inter-module connections was negatively correlated with SARA score, ICARS score, and CAG repeat number after controlling the effects of gender, age and education (SARA: r = -0.56, P = 0.021; ICARS: r = -0.62, P = 0.008; CAG repeat: r = -0.61, P = 0.010; Fig. 3D), but not correlated with disease duration (P = 0.065). The correlations between other network metrics and clinical scores were found to be not statistically significant.

### Relating regional WM network attributes in SCA3 to variation in gene expression patterns

We used partial least square (PLS) regression to relate regional network attributes to gene expression profiles (Fig. 4A). The score of the component in PLS regression, which is the sum of weighted gene expression values, significantly related to regional network attributes and explained about 20% of the variance in the SCA3-related alterations (𝑁𝐸_𝑔𝑙𝑜𝑏_: r = 0.45, P = 0.0166, r^2^ = 0.20, shown in Fig. 4B; 𝑁𝐸_𝑙𝑜𝑐_: r = 0.42, P = 0.0275, r^2^ = 0.17, shown in Fig. 4C; degree centrality: r = 0.48, P = 0.0101, r2 = 0.23). The weights of genes contributing to the component were used to rank the genes. We selected top 30% positive weighted and 30% negative weighted genes and performed gene set enrichment analysis, respectively. The results of GO analysis for top positive weighted genes in 𝑁𝐸_𝑔𝑙𝑜𝑏_ were shown in Table 3 and Fig. 4D. We found that positive weighted genes were most significantly enriched in biological processes including regulation of trans-synaptic signaling, synaptic signaling, and neuron development. The results of GO analysis for top positive weighted genes in 𝑁𝐸_𝑙𝑜𝑐_ were shown in Table 4 and Fig. 4E. We found that positive weighted genes were most significantly enriched in biological processes including RNA metabolic process, chromosome organization, and peptidyl-lysine modification. The biological processes above are subsets of the “macromolecule metabolic process” set, hence we refer to these as a metabolic profile. The results of tissue-type enrichment analysis showed that positive weighted genes were most significantly enriched in tissue types of brain cerebellum and brain cerebellar hemisphere in both measures (Fig. 4F, Fig. 4G).

**Figure 4.**
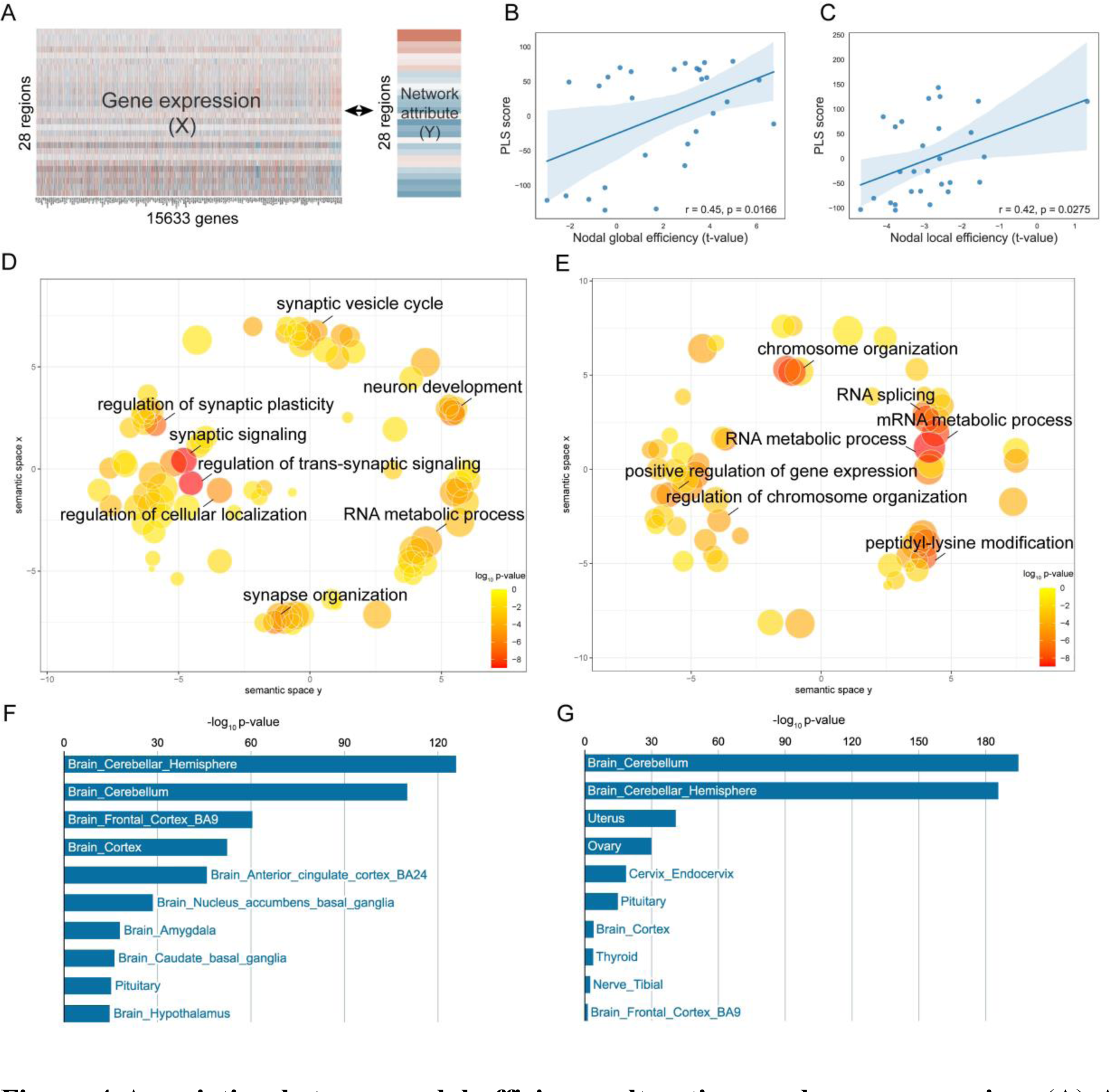
Association between nodal efficiency alterations and gene expression. (**A**) An illustration of bootstrapped PLS regression using gene expression (X) as the predictor variable and network attribute (Y) as the response variable. (**B and C**) Scatterplots show the score of the component in PLS regression significantly related to between-group variances (t-value) of nodal global efficiency and nodal local efficiency. Each dot represents a region. (**D and E**) Gene ontology terms that were significantly enriched in top positive weighted genes are plotted in semantic space, where similar terms are clustered together. (**D)** for nodal global efficiency and (**E)** for nodal local efficiency. Larger, red circles indicate greater significance measured by adjusted p value (see color bar). (**F and G**) Tissue-type enrichment analyses across 54 tissue types obtained from GTEx v8. Top tissue types in the association analysis between gene expression and alterations of nodal attributes are shown, (**F**) for nodal global efficiency and (**G**) for nodal local efficiency.

**Table 3.** Top enriched GO biological processes in positive weighted genes for nodal global efficiency.

| GO term name | N<sup>a</sup> | n<sup>b</sup> | P value | Adjusted P value |
| --- | --- | --- | --- | --- |
| Regulation of trans-synaptic signaling | 432 | 105 | 3.11 × 10<sup>-13</sup> | 1.20 × 10<sup>-9</sup> |
| Synaptic signaling | 704 | 150 | 3.26 × 10<sup>-13</sup> | 1.20 × 10<sup>-9</sup> |
| Regulation of synaptic plasticity | 181 | 55 | 2.01 × 10<sup>-11</sup> | 4.92 × 10<sup>-8</sup> |
| Neuron development | 1082 | 199 | 9.11 × 10<sup>-11</sup> | 1.67 × 10<sup>-7</sup> |
| Synapse organization | 400 | 92 | 2.23 × 10<sup>-10</sup> | 3.28 × 10<sup>-7</sup> |
| Regulation of cellular localization | 872 | 162 | 2.71 × 10<sup>-9</sup> | 3.33 × 10<sup>-6</sup> |
| Cell-cell signaling | 1619 | 268 | 3.66 × 10<sup>-9</sup> | 3.85 × 10<sup>-6</sup> |
| Synaptic vesicle cycle | 189 | 50 | 2.85 × 10<sup>-8</sup> | 2.62 × 10<sup>-5</sup> |
| RNA metabolic process | 1067 | 185 | 4.50 × 10<sup>-8</sup> | 3.68 × 10<sup>-5</sup> |
| Neurogenesis | 1572 | 255 | 5.84 × 10<sup>-8</sup> | 4.29 × 10<sup>-5</sup> |
| Neuron differentiation | 1328 | 221 | 6.49 × 10<sup>-8</sup> | 4.29 × 10<sup>-5</sup> |
| Negative regulation of biosynthetic process | 1517 | 247 | 7.00 × 10<sup>-8</sup> | 4.29 × 10<sup>-5</sup> |
| Synaptic vesicle localization | 162 | 44 | $8.32 \times 10^{-8}$ | $4.69 \times 10^{-5}$ |
| Cell part morphogenesis | 671 | 126 | $8.94 \times 10^{-8}$ | $4.69 \times 10^{-5}$ |
| Vesicle-mediated transport in synapse | 202 | 51 | $1.07 \times 10^{-7}$ | $5.16 \times 10^{-5}$ |
| Positive regulation of synaptic transmission | 169 | 45 | $1.12 \times 10^{-7}$ | $5.16 \times 10^{-5}$ |
| Ribonucleoprotein complex biogenesis | 439 | 90 | $1.24 \times 10^{-7}$ | $5.35 \times 10^{-5}$ |
| Cell projection organization | 1492 | 242 | $1.33 \times 10^{-7}$ | $5.35 \times 10^{-5}$ |
| Ribonucleoprotein complex subunit organization | 244 | 58 | $1.42 \times 10^{-7}$ | $5.35 \times 10^{-5}$ |
| Regulation of synapse structure or activity | 227 | 55 | $1.52 \times 10^{-7}$ | $5.35 \times 10^{-5}$ |
| Protein modification by small protein conjugation or removal | 1078 | 184 | $1.53 \times 10^{-7}$ | $5.35 \times 10^{-5}$ |
| Protein modification by small protein conjugation | 883 | 156 | $1.63 \times 10^{-7}$ | $5.44 \times 10^{-5}$ |
| Chromosome organization | 1167 | 196 | $1.96 \times 10^{-7}$ | $6.25 \times 10^{-5}$ |
| Cell cycle | 1776 | 279 | $2.55 \times 10^{-7}$ | $7.81 \times 10^{-5}$ |
| Synaptic vesicle exocytosis | 120 | 35 | $2.65 \times 10^{-7}$ | $7.81 \times 10^{-5}$ |
| DNA-templated transcription initiation | 244 | 57 | $3.38 \times 10^{-7}$ | $9.54 \times 10^{-5}$ |
| Cellular macromolecule localization | 1866 | 289 | $5.50 \times 10^{-7}$ | $1.50 \times 10^{-4}$ |

<sup>a</sup>The total number of genes in the gene set.
<sup>b</sup>The number of input genes overlapping with the genes in the gene set.

**Table 4.**
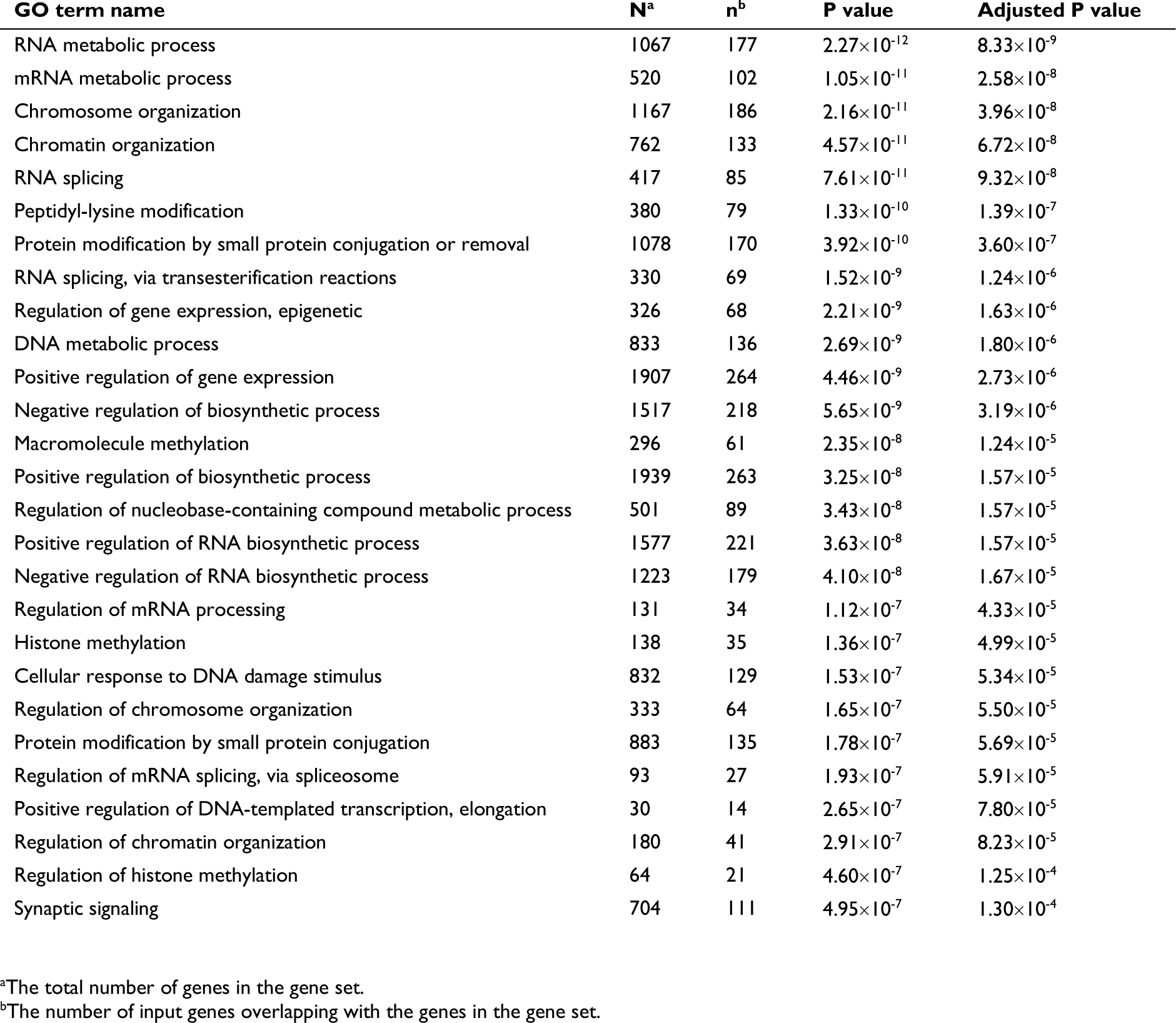
Top enriched GO biological processes in positive weighted genes for nodal local efficiency.

## Discussion

In this study, we investigated topological alterations of cerebellar structural network in patients with SCA3 and for the first time explored their association with gene expression profiles. The decreased global and nodal efficiency, and widely distributed decreased edge strength were observed in SCA3 patients compared with healthy control subjects. Similar hub distributions were found in two groups of subjects, whereas the lower strength of rich-club and feeder connections in SCA3 patients indicated the disruption of hub regions. Moreover, strength of the inter-module connections was lower in SCA3 group and negatively correlated with the CAG repeat number, scores of SARA and ICARS. Finally, transcriptome-connectome association study identified expression of genes involved in synaptic and metabolic processes. Specifically, the variance of regional global efficiency is associated with synaptic genes, whereas the variance of regional local efficiency is associated with metabolic-related genes.

On the global level, SCA3 have less global efficiency, local efficiency, clustering coefficient and more shortest path length compared with controls. The global efficiency and shortest path length both describe the global transmission capability of the network, whereas the local efficiency and clustering coefficient both describe the local transmission capacity of the network^40^. The impaired capacity of both global and local information transmission demonstrated the disruption of both functional integration and segregation in the cerebellum with SCA3, which is consistent with the findings of a previous SCA3 study focused on the WM structural motor network^19^. On the nodal level, the regions with reduced nodal global efficiency were mainly located in bilateral I-IV, bilateral V, bilateral VI, bilateral CrusII, bilateral X and vermis VI. It is noted that bilateral I-IV and bilateral CrusII of them are hub regions of cerebellum in healthy controls. Previously, reduced nodal strength were reported in left I-IV and bilateral V in the preclinical stage of SCA3^19^. The consistent results of areas I-V indicates cerebellar anterior lobe is predominantly affected in SCA3^19^. The regions with reduced nodal local efficiency were mainly located in bilateral VIIIa, right Dentate, bilateral VIIb and right CrusII. In a study of the structural signature in SCA3, volumetric reductions of gray matter were found in similar regions with total regions mentioned above^14^. Remarkably, reduced nodal local efficiency were observed in bilateral Dentate, left Fastigial and left Interposed, which conforms with previous studies that reported cerebellar nuclei as affected neurologic structures in SCA3^6,51^. On the edge level, the widely distributed connections with decreased strength in SCA3 compared with healthy controls indicate the global disruption of structural connectivity in cerebellar network. And the decreased edges were mainly distributed among regions of bilateral I-IV, bilateral V, and their connections with other regions. Previous structural MRI studies reported tissue damage in cerebellar anterior lobe in pre-ataxic group, which was also observed most affected in ataxic stage in our study^19,52^.

Our study has, for the first time, investigated rich-club organization and modular structure in cerebellar structural network. Although the general distribution of hubs is maintained in SCA3, certain hub regions such as cerebellar area I-IV show the most severe disruptions in all network analyses. Moreover, we found that the strength of the rich-club and feeder connections are significantly disrupted in patients, but not for local connections. Combined with the results on the node and edge level, it is concluded that hub regions are most affected in the disease and the affection not only appears among hubs but also extends to the connections between hubs and non-hubs. The pathology of SCA3 preferentially attacks hubs for the reason that their high biological costs lead to more susceptibility to oxidative and metabolic stress^53,54^. The same discovery has been reported in other neurodegenerative disease such as Alzheimer’s disease^55,56^. The modular analysis revealed that the inter-module connections of cerebellum which mostly consist of inter-hemisphere connections significantly disrupted in SCA3. In a previous study, WM loss was found in corticostriatal and inter-hemispheric connections but not for intra-hemispheric connections in Huntington’s disease^57^. It may indicate similar pathological characteristic of WM loss in inter-hemispheric connections for two polyQ diseases. Furthermore, strength of inter-module connections appears to be a potential image biomarker for the disease severity by the negative correlations between the inter-module strengths and scores of SARA and ICARS, and CAG repeat number. As for the correlation between CAG repeat number and imaging parameters, previous studies had controversial findings. A study identified the relationship between the CAG repeat length and brain microstructural alterations of preclinical SCA3^58^, while several studies found no significant correlation between DTI parameters and CAG repeat number^10,11^. Our results supported that the CAG repeat number may intensify white matter damage of cerebellum in SCA3 patients. As for the disease duration, a previous study on white matter motor network in SCA3 reported no significant correlation between network measures and disease duration^19^, which is consistent with our finding.

Our transcriptome-connectome analysis revealed the biological vulnerabilities underlying regional pattern of white matter loss. The genes express higher in regions with more global efficiency decrease in SCA3 were significantly enriched in synapse-related biological processes. And the genes express higher in regions with more local efficiency decrease in SCA3 were significantly enriched in metabolic biological processes. A review article, focused on pathogenesis to therapeutics for SCA3, proposed that misfolding and aggregation of ATXN3 disrupt protein homeostasis and then impact DNA repair, transcription, translation; mitochondria and endoplasmic reticulum; axonal transport and neurotransmission, which lead to dysfunction and disease^59^. It supports our result of enrichment analysis from the way of molecular mechanisms and cellular pathways. Synaptic loss was observed in cerebellum and brainstem of SCA3 patients in PET imaging^60^, and also in other types of hereditary ataxia in the postmortem neuropathology estimation^61,62^. Synapse is responsible for signal passing between neurons, and its abnormality leads to cerebellar neuronal dysfunction and further impacts the output to cerebrum to accurately encode motor-related events^63^. The GO term such as DNA repair is a subset of the term, DNA metabolic process, and RNA splicing is a subset of the term, RNA metabolic process. Previous studies found that ATXN3 interacts with DNA repair proteins^59^, and improperly repaired DNA may activate pro-apoptotic pathways or induce cellular senescence^64–66^. Moreover, CAG repeat can promote aberrant splicing of gene transcript to produce more aggregate-prone disease proteins^67^.

We further discuss the findings of gene expression related to regional global and local efficiency. The global efficiency reflects the information integration of these regions with other regions, so the disruption of integration ability may attribute to abnormal expression of synapse-related genes. Meanwhile, decreased nodal local efficiency which reflects the decline of information segregation may be related to abnormal expression of metabolic genes. Similar findings were reported in a white matter and gene expression association study for Huntington’s disease^68^. The study found that corticostriatal and interhemispheric WM loss is associated with gene expression enriched for synaptic processes, whereas intrahemispheric WM loss is associated with gene expression enriched for metabolic and chromatin-related processes. The interhemispheric connections include more long-range connections which undertake global information transmission and integration, and the intrahemispheric connections undertake more local information transmission and segregation. Collectively, the common findings may imply the biological mechanism of white matter loss in both diseases from integration and segregation perspective.

Several methodological issues should be addressed. First, the gene expression data from AHBA were collected from the brains of six adult donors but only two of them have samples from right hemisphere. We use the data from the whole brain samples, which may cause the data from right hemisphere is not accurate. Moreover, the gene expression data is from six donors who are not diagnosed as SCA3. The transcriptional data sampled from a larger cohort with SCA3 is required for future transcriptome-connectome association study. Second, the sample size is small and the patients vary in disease severity. Future study should recruit more subjects and pick the pre-clinical patients as a dependent group. Finally, besides the cerebellum, the regions such as basal ganglia and thalamus were also reported abnormality in SCA3. These regions can be incorporated into network analysis to focus on their connection to cerebellar regions in further study.

In summary, our study found the disrupted topological organization of cerebellar WM structural connectome in patients with SCA3 and identified expression of genes involved in synaptic and metabolic processes, which suggests a mechanism of white matter regional vulnerability and a potential image biomarker for the disease severity.

## Data Availability

All data produced in the present study are available upon reasonable request to the authors

## Abbreviations

dMRI: diffusion magnetic resonance imaging
ICARS: International Cooperative Ataxia Rating Scale
GO: Gene Ontology
polyQ: polyglutamine
SARA: Scale for the Assessment and Rating of Ataxia
SCA3: spinocerebellar ataxia type 3
SUIT: spatially unbiased atlas template of the cerebellum and brainstem
WM: white matter

